# High proportion of post-acute sequelae of SARS-CoV-2 infection in individuals 1-6 months after illness and association with disease severity in an outpatient telemedicine population

**DOI:** 10.1101/2021.04.24.21256054

**Authors:** James B. O’Keefe, H. Caroline Minton, Colin Johnson, Miranda A. Moore, Ghazala A. D. O’Keefe, Karima Benameur, Jason Higdon, Jessica K. Fairley

**Author notes:** **Corresponding Author:** James B. O’Keefe, Emory Clinic A, 1365 Clifton Road NE Atlanta, GA 30322, USA.

## Abstract

**Background:** Individuals with coronavirus disease 2019 (COVID-19) may have persistent symptoms following their acute illness. The prevalence and predictors of these symptoms, termed post-acute sequelae of SARS-CoV-2 (PASC), are not fully described.

**Methods:** Participants discharged from an outpatient telemedicine program for COVID-19 were emailed a survey (1-6 months after discharge) about ongoing symptoms, acute illness severity, and quality of life. Standardized telemedicine notes from acute illness were used for covariates (comorbidities and provider-assessed symptom severity). Bivariate and multivariable analyses were performed to assess predictors of persistent symptoms.

**Results:** Two hundred and ninety patients completed the survey, of whom 115 (39.7%) reported persistent symptoms including fatigue (n= 59, 20.3%), dyspnea on exertion (n=41, 14.1%), and mental fog (n=39, 13.5%) among others. Proportion of persistent symptoms did not differ based on duration since illness (<90 days: n=32, 37.2% versus ≥90 days: n=80, 40.4%, p = 0.61). Predictors of persistent symptoms included provider-assessed moderate-severe illness (aOR 3.24, 95% CI 1.75, 6.02), female sex (aOR 1.99 95% 0.98, 4.04; >90 days out: aOR 2.24 95% CI 1.01, 4.95), and middle age (aOR 2.08 95% CI 1.07, 4.03). Common symptoms associated with reports of worse physical health included weakness, fatigue, myalgias, and mental fog.

**Conclusions:** Symptoms following acute COVID-19 are common and may be predicted by factors during the acute phase of illness. Fatigue and neuropsychiatric symptoms figured prominently. Select symptoms seem to be particularly associated with perceptions of physical health following COVID-19 and warrant specific attention on future studies of PASC.

## Introduction

In response to the severe acute respiratory syndrome coronavirus 2 (SARS-CoV-2) pandemic, health systems in the United States rapidly reorganized in March 2020 to create home monitoring programs (1). The resulting systems did not anticipate prolonged symptoms following acute coronavirus disease 2019 (COVID-19) until reports arose in personal accounts and social media (2). Subsequent evidence emerged that persistent symptoms are common for both hospitalized (3) and non-hospitalized (4) cohorts, and these patients have described challenges with stigma as well as difficulty accessing care for their symptoms (5). Later reports indicate disruption to usual activities and impaired quality of life may last months (6–8). Studies of survivors of severe acute respiratory syndrome (SARS) hospitalized from 2001-2003 showed persistent pulmonary dysfunction (9) as well as chronic fatigue and psychiatric illnesses (10) lasting years. Other viral illnesses have varied post-acute syndromes including Ebola (11), Chikungunya (12), and dengue (13). Preliminary data for SARS-CoV-2 demonstrate a clear need to characterize the chronic course of this illness. In addition to understanding predictors of chronic COVID-19, understanding the long-term impact on individuals’ health and wellbeing will inform future standards of care for treating patients with COVID-19.

Descriptions of the clinical syndrome of “long COVID” or “post-acute sequelae of SARS-CoV-2 infection” (PASC) have defined symptoms greater than 3 weeks as “post-acute COVID” and greater than 12 weeks as “chronic COVID” (14,15). The underlying pathophysiology is not clear, with studies highlighting possible roles of elevated inflammatory markers, lung dysfunction, muscle weakness, and brain positron emission tomography hypometabolism (16–20). One report using quantitative magnetic resonance imaging could not link specific symptoms to specific organ impairment (21). A meta-analysis of 47,910 patients found the most frequent symptoms include fatigue, headache, attention disorder, hair loss, and dyspnea (22) and noted a high degree of heterogeneity in published studies with a need for future studies to stratify results by patient baseline characteristics and disease severity. Individual cohort studies have reported a variety of specific risk factors for prolonged symptoms including female sex (23–25), higher body mass index (BMI) (23,24), middle age (26), older age (24,27), underlying autoimmunity (8), while one study found no significant predictors (28). Features of acute COVID-19 may predict PASC, with cohort studies finding increasing risk for: patients with more than 5 acute symptoms (24), presence of specific acute symptoms (chest pain, fatigue, fever, olfactory impairment, headaches, or diarrhea) (25), and hospitalization (26).

We previously described duration of symptoms in acute COVID-19 in a telemedicine cohort, and found they were significantly associated with the severity of symptoms at initial medical evaluation (5-6 days after onset) (29). We also noted a higher rate of asthma amongst patients with requiring ongoing telemedicine care for symptoms beyond 6 weeks (42%) compared to the whole cohort (13%) (30). We hypothesize that initial symptom severity as well as select comorbidities such as asthma will also predict prolonged symptoms (>90 days). Lastly, using modified quality of life questions, we explored which persistent symptoms were associated with lower ratings of physical and emotional health.

## Methods

### Study population

Our study population consisted of patients enrolled in Emory Healthcare’s Virtual Outpatient Management Clinic (VOMC) in Atlanta, Georgia between March 24 and September 20, 2020. Patients were offered enrollment in VOMC during results notification calls following a positive SARS-CoV-2 nasopharyngeal reverse transcription polymerase chain reaction (RT-PCR) test at an Emory site (two large throughput outpatient testing centers and the emergency departments of four acute care hospitals). Patients who received a positive test at an outside site (rapid test or RT-PCR) could be referred to VOMC by their Emory provider or by calling the institutional COVID Hotline and hospitalized patients could be referred at discharge if within their isolation period. Patients in VOMC underwent standardized telemedicine assessment by Emory Healthcare providers as previously described (31). Care through VOMC continued for up to 21 days depending on symptom severity and risk factors for hospitalization. Our inclusion criteria were (a) adults at least 18 years of age, (b) previous positive test for SARS-CoV-2, (c) with a valid email address in the Emory electronic health record, and (d) discharge from the Emory VOMC.

### Data collection

A quantitative, cross-sectional survey was created using Qualtrics® Online Survey Software (Qualtrics, Provo, UT) to collect participant-reported information on persistent COVID-19 symptoms and recall of acute symptoms. From August to November of 2020, an email was sent to eligible participants inviting them to complete the survey using an identified link. Non-respondent patients were sent reminder emails and called once by study staff. The survey included research consent, general information (race, sex, education, income level), aspects of the participant’s experience with SARS-CoV-2 infection such as severity of their acute disease, which symptoms were ongoing at the time of the survey (33 individual symptoms), questions related to physical and emotional quality of life, and access to healthcare. Specific symptoms of interest are listed in the results tables. Data on specific comorbidities (see results tables) and provider-assessed severity of acute illness at VOMC intake visit were extracted from the medical record for each participant. Provider-assessed severity was defined as “mild”: cough without shortness of breath or wheeze and / or fever, chills and other mild systemic symptoms; “moderate”: any mild symptoms plus dyspnea on exertion, wheezing and / or mid-chest tightness; “severe”: shortness of breath at rest, pulse oximetry ≤ 92%, pleuritic pain and concerning systemic symptoms like syncope, severe weakness, confusion or acute decline in functional status (29,31). The survey was closed after three months of gathering responses.

### Data analysis

Data from the survey were exported from Qualtrics into Microsoft Excel and analyzed using SAS v9.4 (Carey, NC). A p-value of 0.05 was considered statistically significant for all analyses. Summary statistics were reported using frequencies and means / medians where appropriate. Bivariate analyses using chi-sq, t-test or crude logistic regression (single variable) were performed between the main outcome (persistent symptoms), the main exposure (provider-assessed severity of acute illness) and all covariates. Covariates included age, sex, race, comorbid conditions, hospitalization, and income. Multivariate logistic regression was then performed using any persistent symptom as an outcome with provider-assessed severity (mild versus moderate or severe) as the main exposure and age, sex, hospitalization and comorbidities as confounders. Tests of collinearity and interactions were performed and variables dropped if necessary. Backward stepwise elimination was then undertaken to determine the final model. A second model was conducted limited to participants who answered the survey at 90 days or greater.

In order to determine if certain categories of symptoms had unique associations with potential risk factors for persistence of these symptoms, we divided them into respiratory (dry or wet cough, dyspnea on exertion, shortness of breath at rest, chest tightness or pain), systemic (fatigue, chills, weakness, joint pains, muscle pains, dizziness), neuropsychiatric (headache, mental fog, difficulty sleeping, irritability, feeling depressed, feeling anxious), and loss of taste and/or smell. We then performed bivariate analyses with these outcomes and all main predictors as per the initial analyses above. Also compared to these categories of symptoms were individual rating of severity of acute symptoms related to that specific persistent symptom (like lower respiratory symptoms for persistent respiratory symptoms).

Multivariate logistic regression was performed for each category of symptom using the main exposure of provider rated illness severity and other covariates as listed above. Model diagnostics were also undertaken including testing of collinearity, interactions and confounding. Individual ratings of symptom severity were not included in the multivariate model.

Lastly, all symptoms were compared to measures of physical and emotional health to determine associations with persistent symptoms and three different self-reported markers of quality of physical and emotional health – whether they report their physical health as worse or much worse than before their COVID-19 (versus same or better), whether their physical health limits their daily activities, and whether their emotional health limits their daily activities (yes or no). Bivariate chi square analyses were performed with each symptom except for those persistent symptoms that were reported in less than 5 individuals.

### Ethical Approvals

The study was approved by the Emory University Institutional Review Board. All participants provided electronic consent at the outset of the emailed survey.

## Results

### Basic descriptive factors

Two hundred ninety individuals consented and completed the questionnaire, of whom 216 (75%) were women and 194 (67.4%) reported a comorbid condition (Table 1). The median age was 44 (range 18, 84) and 125 (43.1%) were Black. The median length of time after diagnosis of their acute COVID-19 was 119 days (range 26, 220), with 70% responding at more than 90 days after their acute illness. Of the sample, 35 (12.6%) were hospitalized, and at the initial intake VOMC visit, 182 (68.7%) were determined to have mild or resolved symptoms, 81 (30.1%) moderate, and 2 (0.8%) severe. The most common comorbid condition was obesity (BMI ≥ 30 kg / m^2^) followed by hypertension and asthma / COPD (Table 1). Half of the study sample experienced self-reported weight loss during their illness with a mean loss of 10.7 lbs (SD 7.4) (Table 2). About 25% of individuals said their thinking was worse or much worse than before their acute COVID-19 illness and 25.6% said their physical health was somewhat or much worse than before their acute illness (Table 2).

**Table 1.** Basic demographics and baseline factors for persistent symptoms versus no persistent symptoms at time of survey (ranging 1-7 months post-acute illness). Table 6 has these broken down by early (<90 days) and late (≥ 90 days).

|  | **Persistent symptoms**<br>**n=115** | **No persistent symptoms**<br>**n=175** | **Total**<br>**n=290** |
| --- | --- | --- | --- |
| **Age, years; median (range)** | 46 (20, 84) | 40 (18, 83) | 44 (18, 84) |
| **Sex<sup>i</sup>, n (%)** |  |  |  |
| **Male** | 22 (19.3) | 50 (28.7) | 72 (25) |
| **Female** | 92 (80.7) | 124 (71.3) | 216 (75) |
| **Race / Ethnicity<sup>ii</sup>, n (%)** |  |  |  |
| **White** | 50 (44.6) | 70 (41.4) | 120 (42.7) |
| **Black** | 46 (41.1) | 79 (46.8) | 125 (44.5) |
| **Hispanic / Latinx** | 9 (5.3) | 4 (3.6) | 13 (4.6) |
| **Asian** | 6 (5.4) | 6 (3.6) | 12 (4.3) |
| **Native Hawaiian, Pacific**<br>**Islander** | 0 (0) | 1 (0.6) | 1 (0.4) |
| **Other** | 4 (2.4) | 6 (5.4) | 10 (3.6) |
| **Provider rated moderate –**<br>**severe dx<sup>iii</sup>, n (%)** | 51 (48.1) | 32 (20.1) | 83 (31.3) |
| **Hospitalized<sup>iv</sup>, n (%)** | 20 (18.4) | 15 (8.9) | 35 (12.6) |
| **Any Comorbidity<sup>+</sup>, n (%)** | 76 (67.3) | 118 (67.4) | 194 (67.4) |
| **Lung disease\*** | 24 (23.5) | 25 (16) | 49 (19) |
| **Obesity** | 54 (47.8) | 81 (46.3) | 113 (39.2) |
| **Hypertension** | 27 (26.5) | 47 (30.1) | 74 (28.7) |
| **Heart disease^** | 7 (6.9) | 9 (5.8) | 16 (6.2) |
| **Cancer** | 6 (5.9) | 15 (9.6) | 21 (8.1) |
| **Diabetes** | 11 (10.8) | 14 (9.0) | 25 (9.7) |
| **Immune suppression** | 12 (11.8) | 19 (12.2) | 31 (12) |
| **Kidney disease** | 5 (4.9) | 3 (1.9) | 8 (3.1) |
| **Income<sup>v</sup>** |  |  |  |
| **Less than \$30,000** | 8 (7.5) | 24 (15.4) | 32 (12.2) |
| **\$30,000-\$60,000** | 32 (29.9) | 54 (36.6) | 86 (32.7) |
| **\$60,000 - \$100,000** | 31 (29.0) | 34 (21.8) | 65 (24.7) |
| **Over \$100,000 (ref)** | 36 (33.6) | 44 (28.2) | 80 (30.4) |

\*Defined as asthma, COPD or other chronic lung disease; ^Defined as coronary artery disease or heart failure

**Table 2.** Other characteristics of study survey responses.

| **Other survey variables** | **n (%) unless**<br>**otherwise noted** |
| --- | --- |
| **Weight loss, n (%)** | 132 (50.2) |
| **If yes, average lbs lost (SD)** | 10.7 (7.4) |
| **Sought care for persistent symptoms (n = 113)** |  |
| **No** | 51 (46.0) |
| **Primary Care** | 39 (34.5) |
| **Specialist** | 12 (10.6) |
| **Urgent care** | 1 (.0) |
| **Two or more (i.e. urgent care and primary care)** | 9 (8.0) |
| **Thinking /memory presently** |  |
| **Excellent** | 74 (25.6) |
| **Very good** | 87 (30.1) |
| **Good** | 78 (27.0) |
| **Fair** | 42 (14.5) |
| **Poor** | 8 (2.8) |
| **Compared to before COVID-19, how are thinking &**<br>**memory:** |  |
| **Much better now** | 9 (3.1) |
| **Better now** | 8 (2.8) |
| **About the same** | 198 (68.5) |
| **Worse now** | 69 (23.9) |
| **Much worse now** | 5 (1.7) |
| **Physical health compared to prior to COVID-19 illness** |  |
| **Much better** | 22 (7.7) |
| **Somewhat better** | 35 (12.2) |
| **About the same** | 156 (54.6) |
| **Somewhat worse** | 62 (21.7) |
| **Much worse** | 11 (3.9) |
| **Physical Health affects daily activities** |  |
| **Yes** | 90 (31.4) |
| **Emotional health affects daily activities** |  |
| **Yes** | 83 (29.1) |

### Symptoms

Overall, 115 (39.7%) had persistent symptoms at the time of the survey (Supplemental Table 1). The most common persistent symptom was fatigue (n= 59, 20.3%) followed by dyspnea on exertion (n=41, 14.1%), and mental fog (n=39, 13.5%) (Supplemental Table 1). Table 3 shows the study sample split into two time periods: 26-90 days after the acute COVID-19 phase (“early,” about 1-3 months) and 90-220 days after the acute COVID-19 phase (“late,” over 3 months). There were similar proportions of patients with continued symptoms in the early group (n=34, 38.6%) and the late group (n=79, 39.9%), (p = 0.84). Twenty-nine symptoms are tabulated in table 3 and all of them were very similarly ranked in frequency whether the participant was in the early or late group. Loss of smell was common at 11.9% (n=33) even in the late group (n=21, 10.6%), twice as frequent as loss of taste (n=18, 6.3%). To account for the wide range of days post-COVID-19 that participants answered the survey, t-test analyses were performed for all symptoms based on days post-illness and did not show statistically significant differences based on when they took the survey.

**Table 3.** Descriptive variables stratified by time since acute-COVID-19 illness. P-value shown based on chi-sq or t-test where appropriate, significant values <0.05. Table based on 286 participants as 4 were missing dates of acute illness.

| . | 26-89 (median<br>61) days post | 90-220 (median<br>139) days post | Total<br>n=286 | p-value |
| --- | --- | --- | --- | --- |

|  | **acute COVID-19**<br>**n=88** | **acute COVID-19**<br>**n=198** |  |  |
| --- | --- | --- | --- | --- |
| **Age, mean (SD)** | 46 (16) | 45 (14) | 45 (15) | 0.35 |
| **Female sex** | 67 (76.1) | 147 (74.2) | 214 (74.8) | 0.73 |
| **Race^ (279)** |  |  |  |  |
| White | 41 (47.7) | 79 (40.9) | 120 (43) | 0.26 |
| Black / African American | 33 (38.4) | 90 (46.6) | 123 (44.1) |  |
| Hispanic / Latinx | 3 (3.5) | 10 (5.2) | 13 (4.7) |  |
| Asian | 3 (3.5) | 9 (4.7) | 12 (4.3) |  |
| Native Hawaiian / Pacific Islander | 0 | 1 (0.5) | 1 (0.4) |  |
| Other | 6 (7) | 4 (2.1) | 10 (3.6) |  |
| **Hospitalized** | 9 (10.6) | 25 (13.3) | 34 (12.4) | 0.54 |
| **Provider Defined Disease Severity\*** |  |  |  |  |
| Mild | 56 (67.5) | 126 (69.2) | 182 (68.7) | 0.78 |
| Moderate | 27 (32.5) | 54 (29.7) | 81 (30.6) |  |
| Severe | 0 | 2 (1.1) | 2 (0.8) |  |
| **Persistent symptoms** |  |  |  |  |
| **Any** | 34 (38.6) | 79 (39.9) | 113 (39.5) | 0.84 |
| **Fatigue** | 17 (19.3) | 42 (21.2) | 59 (20.6) | 0.71 |
| **Dyspnea on exertion** | 16 (18.2) | 25 (12.6) | 41 (14.3) | 0.22 |
| **Mental fog** | 12 (13.6) | 27 (13.6) | 39 (13.6) | 1.0 |
| **Difficulty sleeping** | 10 (11.4) | 22 (11.1) | 32 (11.2) | 0.95 |
| **Loss of smell** | 12 (13.6) | 21 (10.6) | 33 (11.5) | 0.46 |
| **Headache** | 8 (9.1) | 23 (11.6) | 31 (10.8) | 0.53 |
| **Dry cough** | 8 (9.1) | 17 (8.6) | 25 (8.7) | 0.89 |
| **Feeling depressed** | 7 (8.0) | 19 (9.6) | 26 (9.1) | 0.66 |
| **Muscle aches** | 7 (8.0) | 17 (8.6) | 24 (8.4) | 0.86 |
| **Joint pains** | 6 (6.8) | 16 (8.1) | 22 (7.7) | 0.71 |
| **Weakness** | 9 (10.2) | 12 (6.1) | 21 (7.3) | 0.21 |
| **Heart palpitations** | 5 (5.7) | 16 (8.1) | 21 (7.3) | 0.47 |
| **Anxiety / nervousness** | 4 (4.6) | 15 (7.6) | 19 (6.6) | 0.34 |
| **Chest tightness / pain** | 6 (6.8) | 13 (6.6) | 19 (6.6) | 0.94 |
| **Loss / change in taste** | 7 (8.0) | 11 (5.6) | 18 (6.3) | 0.44 |
| **Sinus congestion** | 4 (4.6) | 13 (6.6) | 17 (5.9) | 0.51 |
| **Irritability** | 5 (5.7) | 11 (5.7) | 16 (5.7) | 0.97 |
| **Back pain** | 4 (4.6) | 11 (5.6) | 15 (5.2) | 0.72 |
| **Dizziness** | 3 (3.5) | 8 (4.0) | 12 (4.2) | 0.84 |
| **Rhinorrhea** | 2 (2.3) | 9 (4.6) | 11 (3.9) | 0.36 |
| **Ear fullness** | 2 (2.3) | 8 (4.0) | 10 (3.5) | 0.45 |
| **Heartburn** | 2 (2.3) | 8 (4.0) | 10 (3.5) | 0.45 |
| **Shortness of breath at rest** | 2 (2.3) | 7 (3.5) | 9 (3.2) | 0.57 |
| **Nausea** | 1 (1.1) | 4 (2.0) | 5 (1.8) | 0.60 |
| **Rash** | 1 (1.1) | 3 (1.5) | 4 (1.4) | 0.80 |
| **Abdominal pain** | 0 (0) | 4 (2.0) | 4 (1.4) | 0.18 |
| **Chills** | 1 (1.1) | 2 (1.0) | 3 (1.1) | 0.92 |
| **Sore throat** | 1 (1.1) | 1 (0.5) | 2 (0.7) | 0.55 |
| **Diarrhea** | 0 (0) | 2 (1.0) | 2 (0.7) | 0.34 |
| **Loss of appetite** | 1 (1.1) | 0 (0) | 1 (0.4) | -- |
| **Other symptoms** | 3 (3.4) | 11 (5.6) | 14 (4.9) | 0.44 |

^available for 279 participants; \*available for 265 participants

### Predictors of Persistent Symptoms

Since the two time periods (early and late) did not differ in the frequency of persistent symptoms or frequency of specific symptoms, we analyzed the study sample first in aggregate. In addition, we repeated the logistic regression model with only those “late” participants who answered the survey at 90 days or later from acute illness. Our main predicted exposure of moderate to severe disease severity at time of diagnosis (intake visit) was highly associated with persistent symptoms on both unadjusted and adjusted models (Table 4), with an aOR of 3.24 (95% CI 1.75, 6.02). This was similar in the 90 days and greater group. Being hospitalized was also more common in those with persistent symptoms but did not reach statistical significance on the adjusted logistic regression models. In the overall group, female sex was not statistically significantly associated with persistent symptoms, but when the model was limited to the late reporters, female sex (aOR 2.73, 95% CI 1.10, 6.79) was associated. Middle-age (aOR 2.08, 95% CI 1.07, 4.03) was associated with having persistent symptoms in both adjusted models. The only comorbid condition to have a significant result in the model was hypertension which had a negative association with persistent symptoms (aOR 0.45, 95% CI 0.22, 0.94), that remained when limited to the longer duration of symptoms (Table 4).

**Table 4.**
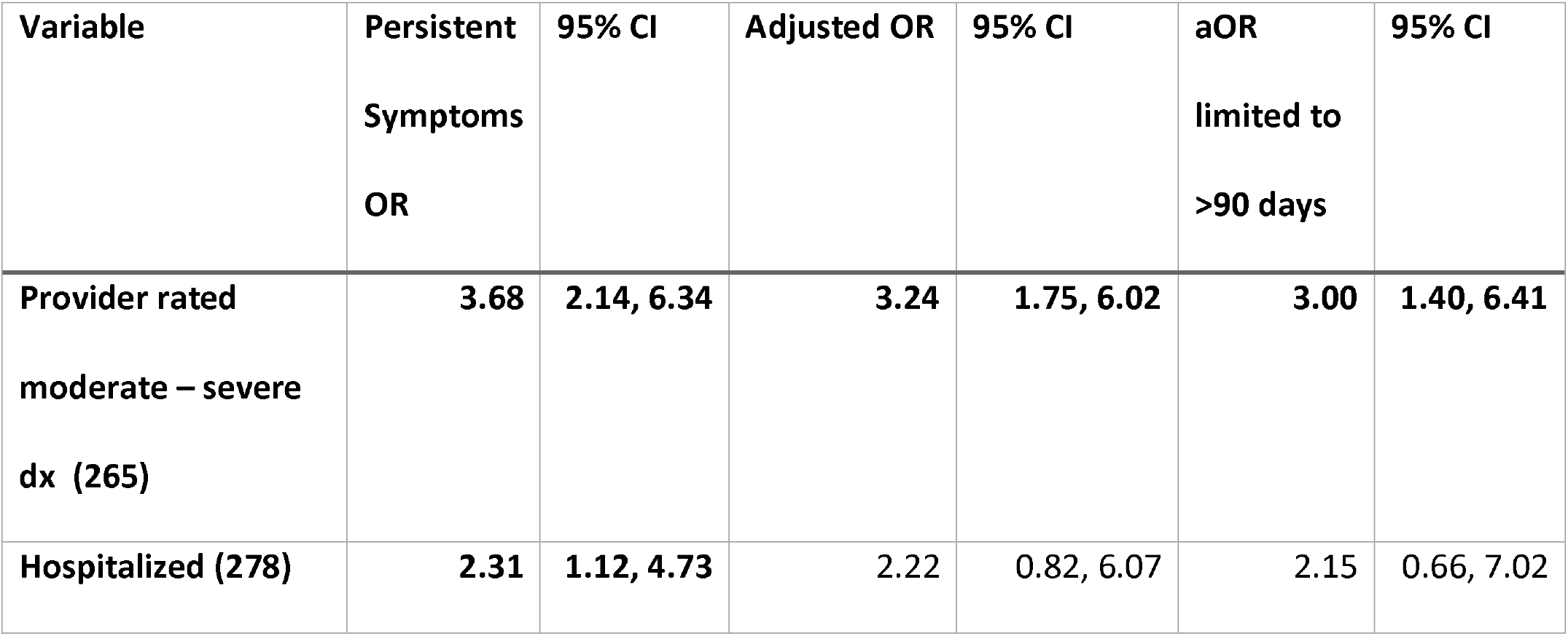
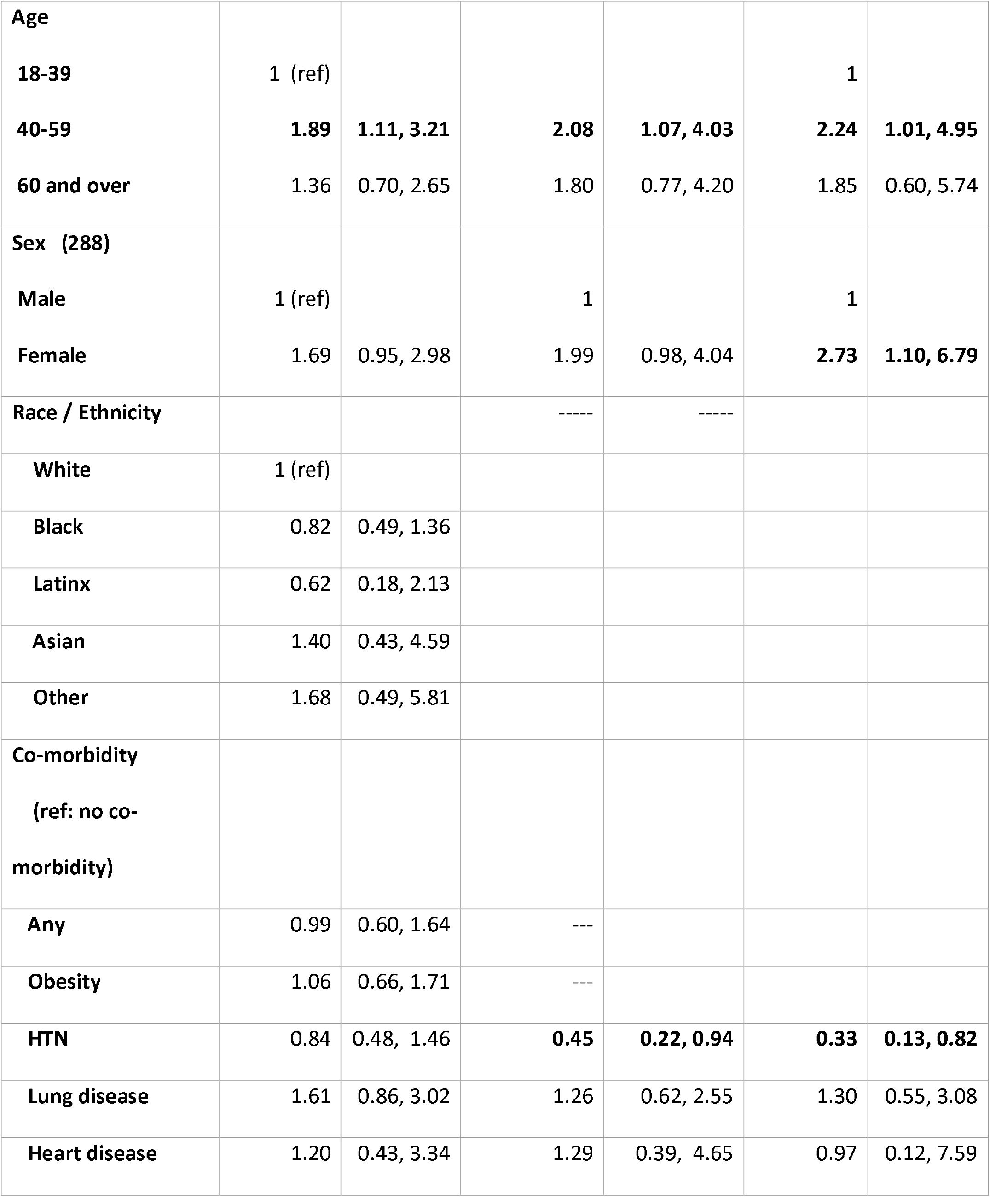
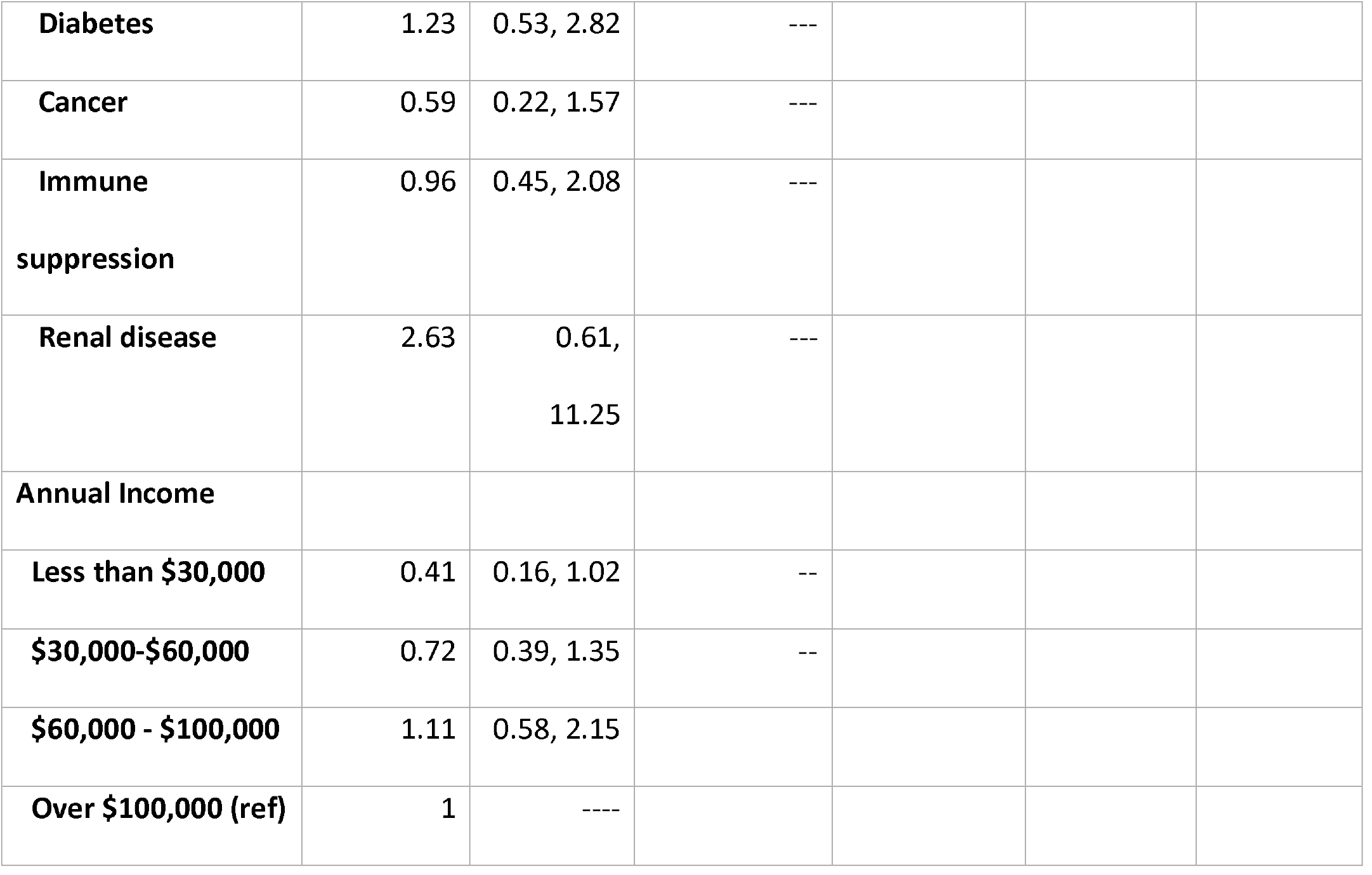
Bivariate analyses and multivariate logistic regression showing unadjusted and adjusted odds ratio respectively for the outcome of persistent symptoms, with moderate-severe acute illness as the main exposure. Bolded values are significant with a p-value of <0.05.

| **Variable** | **Persistent Symptoms OR** | **95% CI** | **Adjusted OR** | **95% CI** | **aOR limited to >90 days** | **95% CI** |
| --- | --- | --- | --- | --- | --- | --- |
| **Provider rated moderate – severe dx (265)** | **3.68** | **2.14, 6.34** | **3.24** | **1.75, 6.02** | **3.00** | **1.40, 6.41** |
| **Hospitalized (278)** | **2.31** | **1.12, 4.73** | 2.22 | 0.82, 6.07 | 2.15 | 0.66, 7.02 |
| **Age** |  |  |  |  |  |  |
| **18-39** | 1 (ref) |  |  |  | 1 |  |
| **40-59** | **1.89** | **1.11, 3.21** | **2.08** | **1.07, 4.03** | **2.24** | **1.01, 4.95** |
| **60 and over** | 1.36 | 0.70, 2.65 | 1.80 | 0.77, 4.20 | 1.85 | 0.60, 5.74 |
| **Sex (288)** |  |  |  |  |  |  |
| **Male** | 1 (ref) |  | 1 |  | 1 |  |
| **Female** | 1.69 | 0.95, 2.98 | 1.99 | 0.98, 4.04 | **2.73** | **1.10, 6.79** |
| **Race / Ethnicity** |  |  | ----- | ----- |  |  |
| **White** | 1 (ref) |  |  |  |  |  |
| **Black** | 0.82 | 0.49, 1.36 |  |  |  |  |
| **Latinx** | 0.62 | 0.18, 2.13 |  |  |  |  |
| **Asian** | 1.40 | 0.43, 4.59 |  |  |  |  |
| **Other** | 1.68 | 0.49, 5.81 |  |  |  |  |
| **Co-morbidity**<br>(ref: no co-morbidity) |  |  |  |  |  |  |
| **Any** | 0.99 | 0.60, 1.64 | --- |  |  |  |
| **Obesity** | 1.06 | 0.66, 1.71 | --- |  |  |  |
| **HTN** | 0.84 | 0.48, 1.46 | **0.45** | **0.22, 0.94** | **0.33** | **0.13, 0.82** |
| **Lung disease** | 1.61 | 0.86, 3.02 | 1.26 | 0.62, 2.55 | 1.30 | 0.55, 3.08 |
| **Heart disease** | 1.20 | 0.43, 3.34 | 1.29 | 0.39, 4.65 | 0.97 | 0.12, 7.59 |
| **Diabetes** | 1.23 | 0.53, 2.82 | --- |  |  |  |
| **Cancer** | 0.59 | 0.22, 1.57 | --- |  |  |  |
| **Immune suppression** | 0.96 | 0.45, 2.08 | --- |  |  |  |
| **Renal disease** | 2.63 | 0.61, 11.25 | --- |  |  |  |
| **Annual Income** |  |  |  |  |  |  |
| **Less than \$30,000** | 0.41 | 0.16, 1.02 | -- | | | |
| **\$30,000-\$60,000** | 0.72 | 0.39, 1.35 | -- | | | |
| **\$60,000 - \$100,000** | 1.11 | 0.58, 2.15 | | | | |
| **Over \$100,000 (ref)** | 1 | ---- | | | | |

When persistent symptoms were divided into categories of symptoms (respiratory, neuropsychiatric, systemic, and loss of taste / smell), the provider-assessed severity at VOMC intake visit remained associated with these persistent symptoms (Table 5) in unadjusted bivariate analyses. Also included were individual patient rating of severity of acute symptoms (recall at time of survey) related to that specific persistent symptom (e.g. patient-rated severity of acute lower respiratory symptoms for persistent respiratory symptoms). Severe ratings for these were also consistently associated with the persistent chronic system-based symptoms.

**Table 5.** Bivariate and adjusted analyses of associated between specific groupings of persistent symptoms broken down by system. Self-rated *acute illness* symptom severity of the same organ symptom also included as a predictor.

|  |  |  |
| --- | --- | --- |
| Symptoms by system | Bivariate associations | Adjusted model |

| **Respiratory Symptoms** | **Persistent**<br>**n=68** | **Not persistent**<br>**n=222** | **OR** | **95% CI** | **aOR** | **95% CI** |
| --- | --- | --- | --- | --- | --- | --- |
| **Provider rated moderate-severe acute illness** | 32 (38.6) | 51 (61.5) | **3.00** | **1.65, 5.78** | **2.85** | **1.43, 5.60** |
| **Hospitalized** | 16 (24.6) | 19 (8.9) | **3.33** | **1.60, 6.95** | **3.17** | **1.20, 8.36** |
| **Self-rated symptom severity during acute COVID-19:** |  |  |  |  |  |  |
| **Lower respiratory** |  |  |  |  |  |  |
| **None or mild** | 10 (14.7) | 117 (52.7) | 1 |  | --- |  |
| **Mod or severe** | 58 (85.3) | 105 (47.3) | **6.46** | **3.14, 13.29** |  |  |
| **Sex** |  |  |  |  |  |  |
| **Male** | 13 (19.1) | 58 (27.4) | 1 |  |  |  |
| **Female** | 55 (80.9) | 154 (72.6) | 1.59 | 0.81, 3.13 | 1.52 | 0.68, 3.37 |
| **Age** |  |  |  |  |  |  |
| **18-39** | 14 (20.6) | 98 (40.5) | 1 | 1 | 1 |  |
| **40-59** | 30 (45.5) | 73 (35.6) | 1.68 | 0.91, 3.12 | 1.37 | 0.65, 2.92 |
| **60 and over** | 12 (18.2) | 34 (16.7) | 1.44 | 0.65, 3.19 | 1.11 | 0.42, 2.92 |
| **Any comorbidity** | 48 (78.7) | 146 (69.9) | 1.59 | 0.81, 3.15 | ----- |  |
| **Lung disease** | 10 (17.5) | 39 (19.4) | 0.88 | 0.41, 1.90 | 0.55 | 0.23, 1.28 |
| **Obesity** | 36 (26.7) | 99 (44.8) | 1.43 | 0.83, 2.48 |  |  |
| **Hypertension** | 21 (36.8) | 52 (25.9) | 1.67 | 0.90, 3.12 | 1.32 | 0.62, 2.81 |
| **Heart disease** | 4 (7.0) | 12 (6.0) | 1.19 | 0.37, 3.84 |  |  |
| **Cancer** | 6 (10.5) | 15 (7.5) | 1.46 | 0.54, 3.95 |  |  |
| **Diabetes** | 8 (14.0) | 17 (8.5) | 1.77 | 0.72, 4.33 |  |  |
| **Immune suppression** | 9 (15.8) | 22 (11.0) | 1.53 | 0.66, 3.53 |  |  |
| **Systemic Symptoms** | **Persistent** | **Not persistent** |  |  |  |  |
|  | **n=74** | **n= 216** |  |  |  |  |
| **Provider rated moderate-severe acute COVID-19 illness** | 37 (52.9) | 46 (23.6) | **3.63** | **2.05, 6.45** | **2.78** | **1.46, 5.32** |
| **Hospitalized** | 14 (19.7) | 21 (10.1) | **2.17** | **1.04, 4.55** | 1.62 | 0.61, 4.33 |
| **Self-rated symptom severity during acute COVID-19:** |  |  |  |  |  |  |
| **General / systemic** |  |  |  |  |  |  |
| **None or mild** | 5 (6.8) | 72 (33.3) | 1 |  | -- |  |
| **Moderate or severe** | 69 (93.2) | 144 (66.7) | **6.90** | **2.67, 17.86** |  |  |
| **Sex** |  |  |  |  |  |  |
| **Male** | 16 (21.6) | 56 (26.2) | 1 |  | 1 |  |
| **Female** | 58 (78.4) | 158 (73.8) | 1.28 | 0.68, 2.42 | 1.27 | 0.59, 2.72 |
| **Age** |  |  |  |  |  |  |
| **18-39** | 23 (31.1) | 103 (48.1) | 1 |  | 1 |  |
| **40-59** | 36 (48.7) | 73 (34.1) | **2.21** | **1.21, 4.04** | 1.98 | 0.96, 4.10 |
| **60 and over** | 15 (20.3) | 38 (17.8) | 1.77 | 0.84, 3.74 | 1.99 | 0.79, 5.02 |
| **Any comorbidity, 288** | 52 (70.3) | 142 (66.4) | 1.20 | 0.68, 2.13 | -- |  |
| **Lung disease, 258** | 19 (28.8) | 30 (15.6) | **2.18** | **1.13, 4.22** | 1.68 | 0.82, 3.47 |
| **Obesity, 288** | 40 (54.1) | 95 (44.4) | 1.47 | 0.87, 2.51 | 1.28 | 0.66, 2.48 |
| **Hypertension, 258** | 20 (30.3) | 53 (27.6) | 1.14 | 0.62, 2.11 | 0.65 | 0.31, 1.40 |
| **Heart disease** | 5 (7.6) | 11 (5.7) | 1.35 | 0.45, 4.04 | --- |  |
| **Cancer** | 4 (6.1) | 17 (8.9) | 0.66 | 0.22, 2.05 | --- |  |
| **Diabetes , 258** | 8 (12.2) | 17 (8.9) | 1.42 | 0.58, 3.46 | --- |  |
| **Immune suppression**<br>**(n=258)** | 10 (15.1) | 21 (10.9) | 1.45 | 0.65, 3.27 | --- |  |
| **Neuropsychiatric Symptoms** | **Persistent**<br>**n=79** | **Not persistent**<br>**n=211** |  |  |  |  |
| **Provider rated moderate-severe illness** | 41 (55.4) | 42 (22) | **4.41** | **2.49, 7.81** | 4.06 | 2.11, 7.81 |
| **Hospitalized** | 17 (22.1) | 18 (9) | **2.90** | **1.40, 5.94** | 3.15 | 1.15, 8.68 |
| **Self-rated symptom severity during acute COVID-19:** |  |  |  |  |  |  |
| **General / systemic** |  |  |  |  |  |  |
| **None or mild** | 7 (8.9) | 70 (33.2) | 1 | -- |  |  |
| **Moderate or severe** | 72 (91.1) | 141 (66.8) | **5.11** | **2.23, 11.68** | ---- |  |
| **Sex** |  |  |  |  |  |  |
| **Male** | 15 (19) | 57 (27.3) | 1 |  | 1 |  |
| **Female** | 64 (81.0) | 152 (72.7) | 1.6 | 0.84, 3.03 | 1.53 | 0.70, 3.36 |
| **Age** |  |  |  |  |  |  |
| **18-39** | 27 (34.2) | 99 (47.4) | 1 |  | 1 |  |
| **40-59** | 37 (46.8) | 72 (34.4) | **1.88** | **1.05, 3.37** | 1.70 | 0.82, 3.51 |
| **60 and over** | 15 (19) | 38 (18.2) | 1.45 | 0.70, 3.01 | 1.36 | 0.52, 3.53 |
| **Any comorbidity** | 55 (69.6) | 139 (66.5) | 1.15 | 0.66, 2.02 | --- |  |
| **Lung disease** | 17 (24.6) | 32 (16.9) | 1.60 | 0.82, 3.12 | 0.96 | 0.45, 2.07 |
| **Obesity** | 43 (54.4) | 92 (44.0) | 1.52 | 0.90, 2.56 | 1.13 | 0.58, 2.20 |
| **Hypertension** | 20 (29) | 54 (28.6) | 1.02 | 0.56, 1.87 | 0.56 | 0.25, 1.28 |
| **Heart disease** | 6 (8.7) | 10 (5.3.) | 1.71 | 0.59, 4.88 | ---- | --- |
| **Cancer** | 5 (7.3) | 16 (8.5) | 0.85 | 0.30, 2.40 | ---- | ---- |
| **Diabetes** | 10 (14.5) | 15 (7.9) | 2.0 | 0.84, 4.61 | 2.20 | 0.74, 6.59 |
| **Immune suppression** | 9 (13) | 22 (11.6) | 1.14 | 0.50, 2.61 | ----- | --- |
| **Loss of taste / smell** | **Persistent** | **Not persistent** |  |  |  |  |
|  | **n=37** | **n=253** |  |  |  |  |
| **Provider rated moderate-severe acute illness** | 17 (48.6) | 66 (28.7) | **2.37** | **1.14, 4.83** | 2.35 | 1.08, 5.11 |
| **Hospitalized** | 2 (5.71) | 33 (13.6) | --- | ----- | ---- | ---- |
| **Self-rated symptom severity during acute COVID-19** |  |  |  |  |  |  |
| **Taste / smell disturbance:** |  |  |  |  |  |  |
| **None or mild** | 1 (2.7) | 99 (39.1) | 1 |  |  |  |
| **Moderate or severe** | 36 (97.3) | 154 (60.8) | 23.1 | 3.12, 171.51 |  |  |
| **Sex** | 30 (80.1) | 186 (74.1) | 1.50 | 0.63, 3.57 | 1.29 | 0.51, 3.26 |
| **Age** |  |  |  |  |  |  |
| **18-39** | 12 (32.4) | 114 (45.4) | 1 |  | 1 |  |
| **40-59** | 20 (54.1) | 89 (35.5) | 2.14 | 0.99, 4.60 | 1.79 | 0.76, 4.21 |
| **60 and over** | 5 (13.5) | 48 (19.1) | 0.99 | 0.33, 2.96 | 1.02 | 0.31, 3.39 |
| **Any comorbidity** | 25 (67.6) | 169 (67.3) | 1.01 | 0.48, 2.11 | -- |  |
| **Lung disease** | 11 (31.4) | 38 (17) | **2.23** | **1.01, 4.94** | 1.90 | 0.82, 4.41 |
| **Obesity** | 18 (48.7) | 117 (46.6) | 1.09 | 0.54, 2.16 | 1.02 | 0.46, 2.26 |
| **Hypertension** | 9 (25.7) | 65 (29.2) | 0.84 | 0.37, 1.89 | 0.72 | 0.28, 1.83 |
| **Heart disease** | 3 (8.6) | 13 (5.8) | 1.51 | 0.41, 5.61 | -- |  |
| **Diabetes** | 3 (8.6) | 22 (9.9) | 0.86 | 0.24, 3.03 | -- |  |
| **Immune suppression** | 5 (14.3) | 26 (11.7) | 1.26 | 0.45, 3.54 | -- |  |

Interestingly, comorbid lung disease was associated with persistent systemic symptoms (OR 2.18, 95% CI 1.13, 4.22) and taste / smell symptoms on bivariate analyses (OR 2.23, 95% CI 1.01, 4.94), but not with persistent respiratory symptoms (0.88, 95% CI 0.41, 1.90). These did not hold in the final logistic models, however. For those that reported moderate to severe taste and smell disturbance during their acute illness (n=190, 65.5%), 81% of those had resolution of their symptoms at the time of their completion of this survey (Table 5).

### Symptoms associated with physical and mental health

Table 6 shows associations with individual symptoms and three different markers of quality of physical and emotional health. For physical health being worse than before acute COVID-19, the commonly reported persistent symptoms most associated were fatigue (OR 10.48, 95% CI 5.47, 20.07), muscle aches (OR 10.47, 95% CI 3.95, 27.81), weakness (OR 10.76, 95% CI 3.75, 30.84), feeling depressed (OR 10.35, 95% CI 4.14, 25.9) and mental fog (OR 9.82, 95% CI 4.63, 20.85. Although less commonly reported and with wide confidence intervals, joint pains (OR 17.1, 95% 5.56, 52.59) and shortness of breath at rest (OR 11.19, 95% CI 2.27, 55.18) had even stronger associations. For physical health limiting daily activities, the common symptoms most associated were dyspnea on exertion (OR 13.68, 95% CI 5.98,31.28), mental fog (OR 12.41, 95% CI 5.41, 28.48), headache (OR 12.24, 95% CI 4.81, 31.17), and joint pains (OR 12.06, 95% CI3.95, 36.85). While not a commonly reported symptom, heart palpitations (OR 16.17, 95% 4.62, 56.53) was strongly associated with limitations of daily activities. For emotional health limiting daily activities, the following symptoms were most associated: irritability (OR 6.02, 95% CI 2.02, 17.9), feeling depressed (OR 5.53, 95% CI 2.35, 12.99), back pain (OR 5.40, 95% CI 1.78, 16.32), feeling anxious (OR 4.71, 95% CI 1.78. 12.43), and ear fullness (OR 3.86, 95% 1.06, 14.04).

**Table 6.** Associations, measured by odds ratios, and 95% confidence individuals, between individual persistent symptoms and physical and emotional health indicators. Limited to symptoms that were reported by at least 5 participants.

| **Persistent symptoms** | **Physical health worse than**<br>**before acute COVID-19** | **Physical health**<br>**affects daily activities** | **Emotional health**<br>**affects daily activities** |
| --- | --- | --- | --- |
|  | OR (95% CI) | OR (95% CI) | OR (95% CI) |
| **Fatigue** | 10.48 (5.47, 20.07) | 10.35 (5.36, 19.98) | 2.56 (1.41, 4.64) |
| **Dyspnea on exertion** | 8.34 (4.06, 17.15) | 13.68 (5.98, 31.28) | 2.16 (1.09, 4.25) |
| **Mental fog** | 9.83 (4.63, 20.85) | 12.41 (5.41, 28.48) | 3.06 (1.53, 6.10) |
| **Difficulty sleeping** | 7.42 (3.36, 16.35) | 7.17 (3.16, 16.27) | 3.21 (1.52, 6.79) |
| **Loss of smell** | 4.32 (2.05, 9.12) | 4.69 (2.19, 10.05) | 1.46 (0.68, 3.12) |
| **Headache** | 3.99 (1.84, 8.66) | 12.24 (4.81, 31.17) | 3.46 (1.62, 7.40) |
| **Dry cough** | 2.34 (1.02, 5.36) | 2.01 (0.89, 4.54) | 1.09 (0.45, 2.62) |
| **Feeling depressed** | 10.35 (4.14, 25.9) | 7.26 (2.93, 18.02) | 5.53 (2.35, 12.99) |
| **Muscle aches** | 10.47 (3.95, 27.81) | 6.32 (2.52, 15.87) | 2.68 (1.15, 6.23) |
| **Joint pains** | 17.1 (5.56, 52.59) | 12.06 (3.95, 36.85) | 1.77 (0.73, 4.31) |
| **Weakness** | 10.76 (3.75, 30.84) | 11.24 (3.66, 34.51) | 1.55 (0.62, 3.89) |
| **Heart palpitations** | 4.46 (1.79, 11.08) | 16.17 (4.62, 56.53) | 2.38 (0.97, 5.84) |
| **Anxiety / nervousness** | 3.60 (1.40, 9.25) | 7.07 (2.46, 20.32) | 4.71 (1.78, 12.43) |
| **Chest tightness / pain** | 7.48 (2.73, 20.51) | 5.38 (1.97, 14.65) | 1.85 (0.72, 4.79) |
| **Loss / change in taste** | 4.67 (1.71, 12.78) | 2.95 (1.12, 7.76) | 1.60 (0.60, 4.28) |
| **Sinus congestion** | 2.79 (1.03, 7.53) | 5.91 (2.01, 17.32) | 1.77 (0.65, 4.82) |
| **Irritability** | 3.15 (1.14, 8.74) | 7.42 (2.32, 23.72) | 6.02 (2.02, 17.9) |
| **Back pain** | 4.85 (1.66, 14.15) | 9.95 (2.73, 36.22) | 5.40 (1.78, 16.32) |
| **Dizziness** | 6.43 (1.88, 22.05) | 7.19 (1.90, 27.22) | 3.63 (1.12, 11.78) |
| **Rhinorrhea** | 3.73 (1.10, 12.60) | 6.31 (1.63, 24.37) | 2.09 (0.62, 7.06) |
| **Ear fullness** | 4.68 (1.28, 17.08) | 9.51 (1.98, 45.76) | 3.86 (1.06, 14.04) |
| **Heartburn** | 3.06 (0.86, 10.89) | 5.45 (1.38, 21.61) | 2.53 (0.71, 8.97) |
| **Shortness of breath at rest** | 11.19 (2.27, 55.18) | 4.62 (1.13, 18.91) | 1.23 (0.30, 5.02) |

## Discussion

### Principal Findings

We find that chronic symptoms are common in a follow-up survey of patients previously enrolled in a COVID-19 outpatient telemedicine monitoring program, with 40% of patients reporting persistence of at least one symptom. We hypothesized based on prior work (29) that initial symptom severity (at time of telemedicine intake visit) would predict the likelihood of long-term symptoms and we find this association is significant. Patient recall of symptom severity (reported at time of survey) finds similar relation strengthening the internal validity of this finding. We do not find association between other factors such as demographics or comorbidities and the presence of symptoms at the time of follow-up survey except for female sex (increased likelihood) and hypertension (decreased likelihood), nor do we find a significant difference between patients responding to the survey closer to their acute illness versus those farther out from their acute illness.

The mechanism(s) underlying the persistence of symptoms beyond acute COVID-19 remain unclear; the finding of a relationship between the severity of acute symptoms (recorded at the end of the first week of illness) and the prevalence of symptoms months later suggests a possible role for acute inflammation and/or viral burden. Subsequent hospitalization may predict risk in the unadjusted comparison but is not significant in the adjusted model. This may be due to the low number of hospitalizations in our cohort, but also that risk factors for hospitalization (age, male sex, comorbidities) are not directly associated with risk of long-term symptoms.

These findings are significant in public health messaging and individual risk communication for patients with COVID-19. The factors predicting persistent symptoms remain unclear and it appears that patients and the public cannot be reassured that individuals are unlikely to develop persistent symptoms until their initial course (i.e. first week of illness) is past. Furthermore, we do not find a difference in the frequency of persistent symptoms in our “early” (day 26-89) and “late” (day 90-220) groups. While it is possible that there are differences between these groups based on the “wave” of the pandemic, it suggests that there is not a clear time for expected resolution of symptoms that persist, and further study will be needed to determine long-term outcomes.

Many respondents to the survey reported lower quality of life on questions regarding limitations due to physical and emotional health, similar to a prior report indicating disrupted home, work, and social life for patients after acute COVID-19(6). These responses were associated with certain common symptoms (e.g. fatigue, dyspnea on exertion, mental fog) more than others (e.g. cough, change in smell or taste). These findings may suggest that specific symptom profiles are more significant in quality of life and may warrant particular emphasis in studies of PASC. More than half sought care for their symptoms indicating a level of concern leading to medical attention. Prior work indicates that patients encounter challenges due to limited recognition of PASC (5,32) and providers should be aware of the likelihood of PASC and evolving treatment guidelines.

### Comparison to prior studies

Similar to previous studies (23,25–27,33,34), this study represents a single institution report of persistent symptoms in patients after acute COVID-19 and finds a high rate of persistent symptoms. A novel aspect of this work is the prospective enrollment in the telemedicine program with standardized data collection by providers in the acute phase of illness, such that risk factors for later outcomes could be systematically analyzed. We found that the severity of symptoms at the time of the acute telemedicine visit was a significant predictor, which corroborates the prior reports relating acute illness profile to prolonged symptoms (24–26). We do not find a strong association with hospitalization, suggesting that the term “severity” may need further clarification as a symptom descriptor distinct from “disease severity” (often referring to highest level of care, which is more strongly associated with age, sex, and medical comorbidity). The use of the physical health questions also gives more description and nuance to our findings, which is an added strength over large cohort studies of reported frequency of persistent symptoms.

We find that female gender is associated with PASC in our cohort, especially in the later group, similar to prior studies (23–25). It is well described that males are at higher risk for severe acute COVID-19 (35), and it has been proposed that genetic differences (including expression of Toll-like receptor 7 on the X chromosome) may account for this(36), but a mechanism relating to increased risk for PASC is not clear. There may be a common mechanism with other post-infectious syndromes: female gender has been reported as associated with lower six minute walk distance in survivors of SARS (9) as well higher rate of post-dengue fatigue (13), post-chikungunya arthralgia (12) but not consistently for post-Ebola symptoms(11).

Middle age (40-59) was significant in the adjusted model, similar to prior report (26). Given the strong association of illness severity (i.e. risk of hospitalization and death) with age(35), hypothesized to relate to immune system changes across age groups(36), it is plausible that immune-system mediated factors during acute illness increase the risk for PASC in individuals in middle age more than other age categories.

The role of hypertension in acute COVID-19 is not clear - initially reported as a possible risk factor for severe COVID-19 (in excess in severe versus nonsevere cases)(37), has not been shown to be a robust predictor of severity with age-hypertension interactions even decreasing risk(35) and improved blood pressure control correlating to worse outcomes (38). Antihypertensive medications may impact disease course, with preliminary results indicating effects that may vary by race(39). In this context, it is possible that there is a blood pressure (or antihypertensive) correlation with PASC and this relationship merits further study.

## Limitations

This report was conducted within a specific cohort (outpatient telemedicine clinic in Atlanta, Georgia) early in the COVID-19 pandemic. To enter this cohort, patients must have had symptoms consistent with COVID-19 and consented to telemedicine visit – few asymptomatic (or minimally symptomatic) patients are represented and the number of hospitalized patients is relatively small, thus limiting generalizability. A related concern is response bias, with a relatively low response rate to the survey. However, we did not indicate the purpose of the survey in the invitation email and consent and do not have reason to believe respondents would be more symptomatic than non-respondents. A third consideration interpreting our results is the uncertainty regarding the definition and grading of PASC. This was not a known entity at the time of survey design and the questions addressed the presence of ongoing symptoms and overall health-related quality of life, but not the impact of individual symptoms on functional status. For this reason, we have analyzed the association of symptoms with functional limitations (Table 5) as a prompt for further study.

Future study is needed to clarify the risk factors, mechanisms, duration, and subtypes of PASC. This preliminary work indicates that there are features of acute COVID-19 that may be predictive and prospective study including objective measure (e.g. inflammatory markers) as well as patient-reported measures (e.g. symptom severity) may clarify this further. We find a high rate of PASC in an outpatient cohort with impact on many individuals quality of life and ongoing follow-up for this important entity is warranted.

## Data Availability

Deidentified data available upon reasonable request to the corresponding author.

## MANUSCRIPT INFORMATION

### Funding

This work was supported by the Georgia Geriatrics Workforce Enhancement Program COVID-19 Telehealth award, which is supported by the Health Resources and Services Administration (HRSA) of the US Department of Health and Human Services (HHS) as part of award number T1MHP39056 totaling US $90,625, with 0% financed with non¬governmental sources. The contents of this paper are those of the authors and do not necessarily represent the official views of, nor an endorsement by, the HRSA, the HHS, or the US Government.

## Acknowledgments

We would like to acknowledge Elizabeth Harrell for data retrieval for this work. We also acknowledge the members of the Virtual Outpatient Management Clinic at Emory Healthcare, including faculty, staff, and administrative members of the Paul W Seavey Comprehensive Internal Medicine Clinic and the Emory at Rockbridge Primary Care clinic, as well as the physicians, nurses, and APPs who volunteered from other sites.

