## Supplemental Table 1 for "High proportion of post-acute sequelae of SARS-CoV-2 infection in individuals 1-6 months after illness and association with disease severity in an outpatient telemedicine population"

Supplemental Table 1. Total numbers and proportion of each persistent symptom in the overall study sample.

| Persistent symptoms | n, (%)  Total = 290 |
| --- | --- |
| Any | 115 (39.7) |
| Fatigue | 59 (20.3) |
| Dyspnea on exertion | 41 (14.1) |
| Mental fog | 39 (13.5) |
| Difficulty sleeping | 32 (11.0) |
| Loss of smell | 33 (11.4) |
| Headache | 31 (10.7) |
| Dry cough | 26 (9.0) |
| Feeling depressed | 26 (9.0) |
| Muscle aches | 24 (8.3) |
| Joint pains | 22 (7.6) |
| Weakness | 21 (7.2) |
| Heart palpitations | 21 (7.2) |
| Anxiety / nervousness | 19 (6.6) |
| Chest tightness / pain | 19 (6.6) |
| Loss / change in taste | 18 (6.2) |
| Sinus congestion | 17 (5.9) |
| Irritability | 16 (5.5) |
| Back pain | 15 (5.2) |
| Dizziness | 12 (4.1) |
| Rhinorrhea | 11 (3.8) |
| Ear fullness | 10 (3.5) |
| Heartburn | 10 (3.5) |
| Shortness of breath at rest | 9 (3.1) |
| Nausea | 5 (1.7) |
| Rash | 4 (1.4) |
| Abdominal pain | 4 (1.4) |
| Chills | 3 (1.0) |
| Sore throat | 2 (0.7) |
| Diarrhea | 2 (0.7) |
| Loss of appetite | 1 (0.3) |
| Other symptoms | 14 (4.8) |
